# Factors Associated with Access and Use of PPE during COVID-19: A Cross-sectional Study of Italian Physicians

**DOI:** 10.1101/2020.04.24.20073924

**Authors:** E Savoia, G Argentini, D Gori, E Neri, R Piltch-Loeb, MP Fantini

**Affiliations:** Emergency Preparedness Research, Evaluation, & Practice (EPREP) program, Division of Policy Translation & Leadership Development, Harvard T.H. Chan School of Public Health, 677 Huntington Avenue, 02115 Boston (MA), USA; IRCCS Burlo Garofolo, Via dell’Istria, 65, 34137 Trieste, Italy; Department of Biomedical and Neuromotor Sciences (DIBINEM), University of Bologna, via San Giacomo 12, 40126, Bologna, Italy; Azienda Sanitaria Giuliano Isontina, Via Costantino Constantinides 2, 34128 Trieste, Italy

## Abstract

**Objectives:** During the course of the Novel Coronavirus (SARS-CoV-2) pandemic, Italy has reported one of the highest number of infections. Nearly ten percent of reported coronavirus infections in Italy occurred in healthcare workers. This study aimed to understand physicians’ access to personal protective equipment (PPE) and to information about their use, risk perception and strategies adopted to prevent contracting the infection.

**Methods:** We undertook a cross-sectional, online self-reported survey implemented between March 31 and April 5 2020 of Italian physicians.

**Results:** Responses were received from 529 physicians, only 13% of which reported to have access to PPE every time they need them. Approximately half of the physicians reported that the information received about the use of PPE was either clear (47%) or complete (54%). Risk perception about contracting the infection was influenced by receiving adequate information on the use of PPE. Access to adequate information on the use of PPE was associated with better ability to perform donning and doffing procedures [OR=2.2 95% C.I. 1.7–2.8] and reduced perception of risk [OR=0.5, 95% C.I. 0.4–0.6].

**Conclusions:** Results from this rapid survey indicate that while ramping up supplies on PPE for healthcare workers is certainly of mandatory importance, adequate training and clear instructions are just as important.

## Introduction

Globally, as the Novel Coronavirus (SARS-CoV-2) pandemic has evolved there has been a shortage of personal protective equipment (PPE) available to the healthcare workforce.^1 2^ As the World Health Organization has warned since the beginning of March, disruption to the global supply of PPE, has left frontline healthcare workers ill-equipped to care for their patients.^2 3^ Since the start of the epidemic, guidance on the usage of such equipment has continued to evolve, and has emphasized conservation of resources rather than optimizing protection of workers.^4^

The coronavirus pandemic has taken a dramatic toll worldwide and especially in Italy. As of the beginning of April, Italy has reported one of the highest number of infections and the highest number of deaths of any European country.^5^ Media reports from across Italy have shone a light on the burden that the coronavirus is placing on health workers. Nearly ten percent of reported coronavirus infections in Italy occurred in healthcare workers.^6 7^ As of April 14, 162.488 cases and 21,067 deaths attributed to COVID-19 were confirmed in the country, and the number of healthcare workers infected and those that lost their life due to COVID-19 was 14,066 and 133 respectively. Many of these infections are likely due to occupational hazard; workers becoming infected while caring for patients suggesting the shortage or inappropriate use of PPE may be at the root of part of these infections.

The use of PPE has been identified as one of the biggest physical and psychological challenges experienced by physicians while responding to COVID-19.^7^ For example, physical burdens related to PPE include repeated donning and doffing of equipment and extended hours wearing uncomfortable masks and respirators, while psychological burdens include challenges communicating with peers and patients when wearing PPE on and operating under changed practice standards. Because of PPE shortages, healthcare workers, who may have been trained on how to don and doff PPE to maximize protection from infection, have had to make ad hoc adjustments on what piece of equipment to use and when, that are not reflected in any training they have received. The additional burdens created by a shortage whereby processes for using PPE are continuously changing, has not been explored.

The Italian healthcare system is regionally based and organized at the national, regional, and local levels, with each region having the autonomy of managing the delivery of the healthcare services based on local needs.^8^ Italian National Health Service system certifies healthcare workers and requires continuing education and quality and standards of care are set by the regions and hospitals. Training procedures for the healthcare workforce are also left to the regions and local hospitals, specifically regarding the management of PPE. Such differences are expected given local needs and hospital settings differ by localities, however such differences may also have caused inconsistencies and confusion on the appropriate use of PPE in a rapidly evolving situation such as the COVID-19 outbreak. Currently, there is lack of literature on how the healthcare workforce in Italy has adapted during the Novel Coronavirus pandemic in the use of equipment. This study aimed to understand physicians’ access to PPE, reception of information about their use, ability to perform donning and doffing procedures, risk perception and strategies adopted to prevent contracting the infection. We believe the results of our work may be helpful in the development of policies and training related to the use of PPE in Italy as well as in other countries.

## Methods

### Study population

We undertook a cross-sectional, self-reported survey, of physicians working in Italy during the response to COVID-19. We disseminated an online survey by the use of two social media groups (via Facebook and WhatsApp) created by physicians engaged in the response. We obtained an institutional review board approval to conduct this study by Harvard Longwood Medical’s Institutional Review Board. The survey was implemented between March 31 and April 5 2020. The population of interest included physicians aged ≥ 21 years with a valid medical license (criteria to join the social media group) and working in Italy during the emergency.

### Survey instrument

The questionnaire was developed through a series of meetings between the researchers and practitioners in charge of infectious control procedures and PPE training activities at the hospital level, the practitioners provided feedback on the content validity and comprehensiveness of the survey instrument before implementation. Questions were designed to inform the development of training and policies in response to the crisis and included questions about the physician’s work experience (years of experience, specialty, experience in COVID-19 units and geographic area of work), and questions related to the use of PPE divided in four parts: 1) Access to PPE and strategies to cope with shortage, 2) Information received on the use of PPE, 3) Self-reported ability to perform donning and doffing procedures, and 4) Risk perception of contracting the disease.

### Data analysis

Our analysis examines four dependent variables: 1) access to PPE, 2) use of PPE, 3) self-reported ability to perform donning and doffing procedures and 4) risk perception in the work setting. To determine the first of these, we asked: 1) *Do you believe to have adequate access to the PPE necessary for your daily professional activity?* Response options were coded as follows: rarely and never =1, sometimes=2, and always=3. For the second dependent variable we asked: 2) *Do you believe you have received adequate information regarding the use of PPE to protect yourself from contracting COVID-19?* Response options were coded as follows: rarely and never =1, sometimes=2, and always=3. For the third dependent variable we asked: *3) Based on the information received to date do you believe you can correctly don and doff the following pieces of equipment* ( a list was provided including respirators, masks, gowns, cups and gloves). Response options were coded as follows: (0=I do not know how to don or doff the piece of equipment, 1= I am not sure, 2=I know how to do it). We combined all responses for each piece of equipment into a scoring system to create a new variable named “*ability to perform donning and doffing procedures”*, we then dichotomized the variable into high ability when the score was 75th percentile and less than high when below the 75th percentile). Finally, to measure risk perception we asked 4) *What do you believe is your risk of contracting COVID-19 in the work setting in the next 30 days?* physicians rated their perception using a scale ranging from 0=no risk to 100=high risk, responses were coded as follows: 1= low risk (≤ 25th percentile), 2=medium risk (26th-85th percentile) and 3=high risk (≥ 75th percentile). We first performed descriptive statistics for each variable. We then applied ordered logistic regression to the three ordinal variables access to PPE, information on PPE use, and donning and doffing ability and logistic regression to the variable risk perception. We tested for bivariate associations between each predictor (years of experience, geographic region, type of position, working in a dedicated COVID-19 unit) and the dependent variables using a p-value ≤0.25 as cut-off as inclusion criteria for the multiple regression model. We tested the parallel regression assumption by means of the *Brant* test for the ordered logistic model which resulted not statistically significant and as such the *ologit* command was used to run the analysis. Qualitative analysis was conducted on open questions to identify how physicians coped with PPE shortages and what strategies they adopted to reduce the risk of infecting their family members. We used Stata 16 software (Stata, College Station, TX) to analyse the data.

## Results

### Characteristics of the study population

Responses were received from 529 physicians, the majority of respondents were in the age category 36–50 (40%), working in the hospital setting as employees of the national healthcare system (59%), and the most frequently reported category for years of experience was 11–20 (30%). Physicians from all 20 Italian regions and the Republic of San Marino were included in our survey, most respondents were from the Lombardia region (13%), the most impacted by the emergency. Over 40 medical specialities were reported by the respondents, the most frequent of which being Pediatrics (12%), Primary care (7%) and Anesthesiology/Intensive Care (6%) and Cardiology (6%). Details on the sample characteristics are provided in Table 1.

**Table 1.**
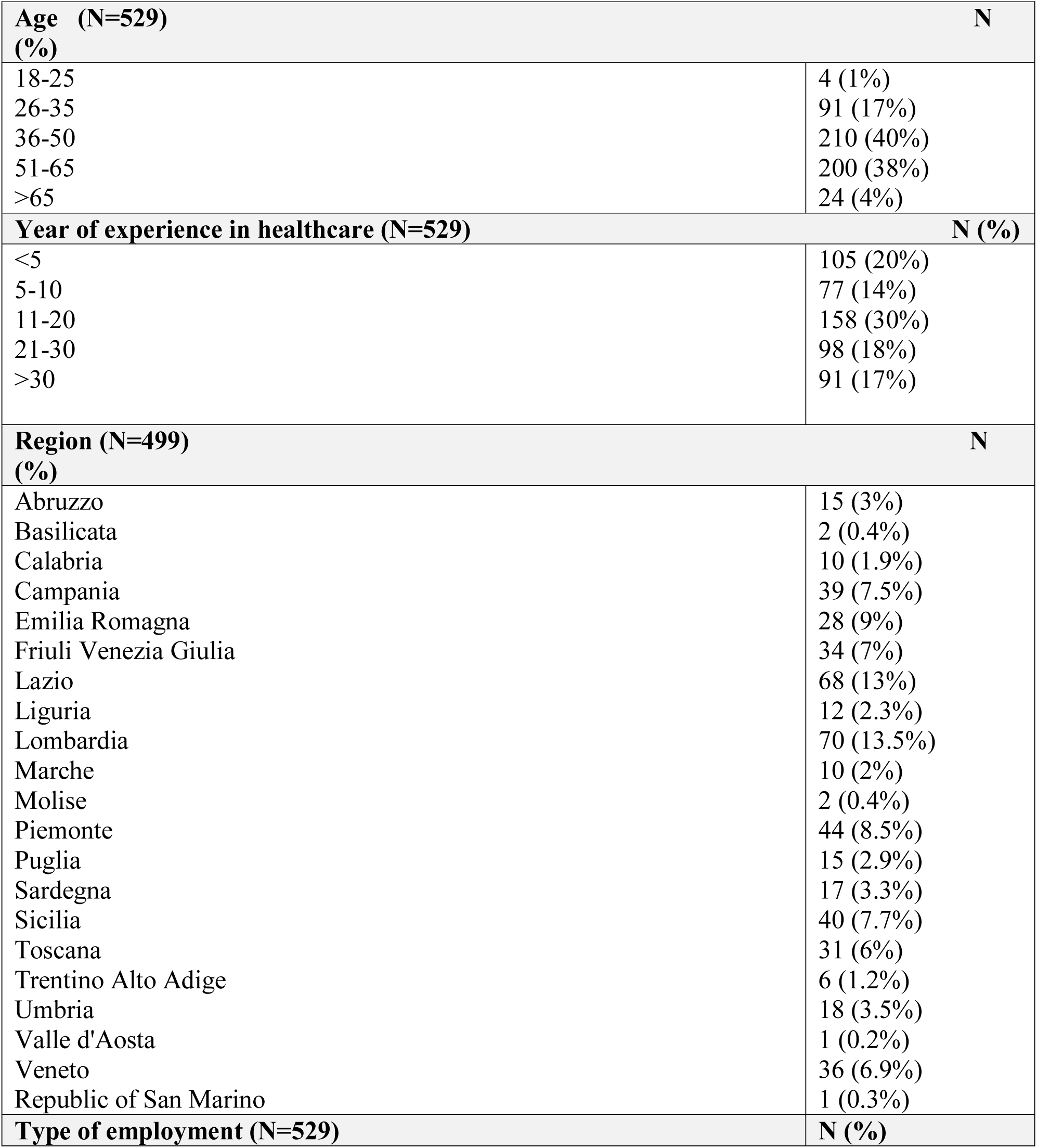
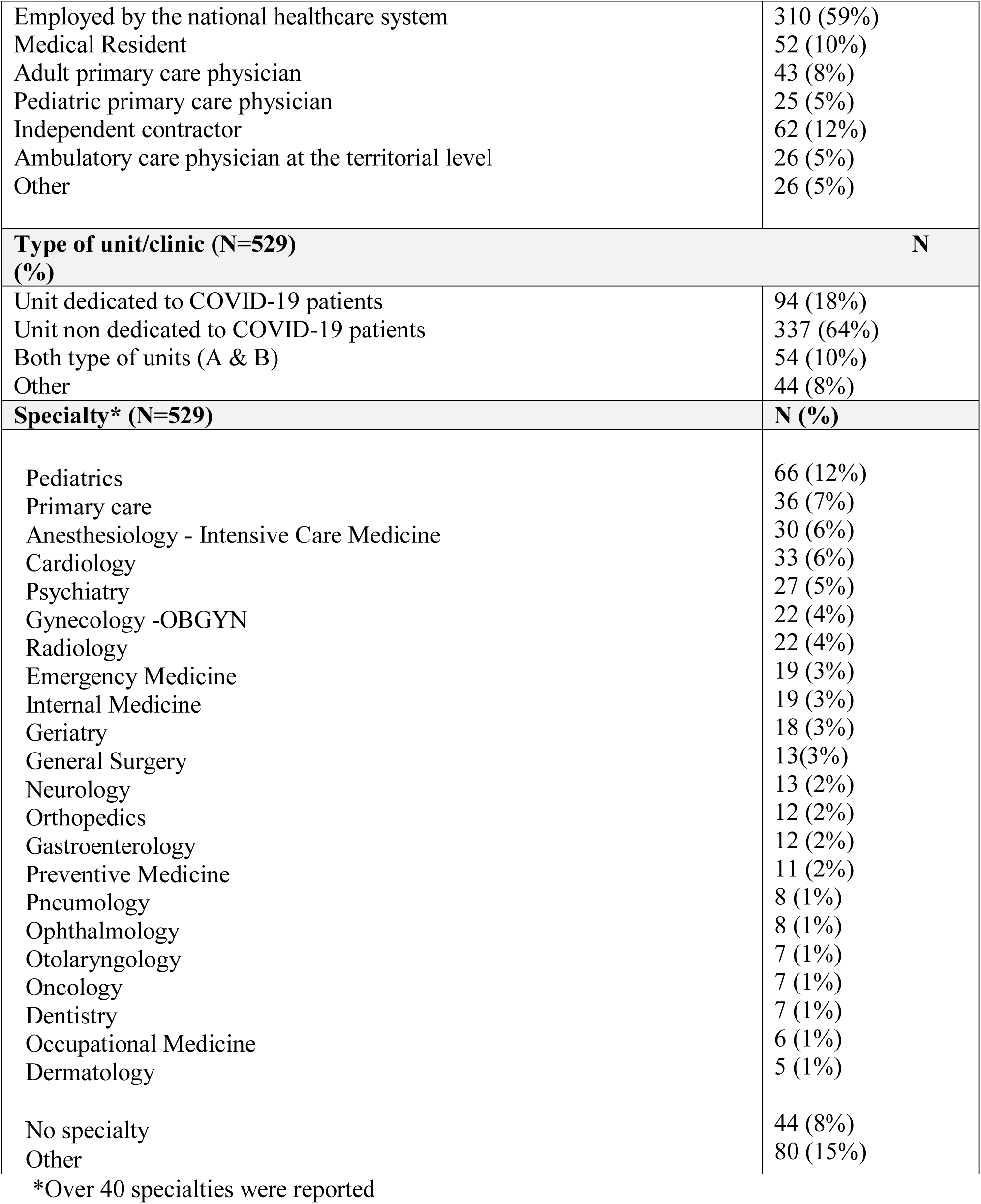
Respondents’ characteristics

### Access to PPE

When asked if they had access to PPE when they needed it, 195 (37%) of the physicians said they rarely or never did, 265 (50%) sometimes and 69 (13%) always did. FFP3 and FFP2 (equivalent to N-99 and N-95 in the USA) were the pieces of equipment most frequently reported as lacking by 59% and 56% of physicians respectively. Other pieces of equipment were also reported as lacking but by a lower percentage of respondents: gown (45%), hair cups (34%), surgical masks (27%), gloves (16%). Lack of PPE forced 89% of physicians to come up with strategies to cope with the shortage. Such strategies included using the same N-95 for long shifts (12 hours and beyond), disinfecting the respirator with alcohol, adding a surgical mask either under or on top of the N-95, re-using the same mask for multiple shifts, exposing the respirator to “the sun” as reported to some of them or ozone, making masks on their own at home, or buying respirators of unknown certification.

In the bivariate analysis of factors that related to PPE access; working in a COVID-19 unit, in the North or Centre of the country and in a primary care setting were associated with access to PPE, while the variable *years of work experience* was dropped from the final model because of p-value > 0.25. More specifically, in the final ordered multiple logistic model physicians working in COVID-19 units had 3.8 increased odds (OR=3.8, 95% C.I. 2.5–5.7) of having *access to the PPE they need* at a higher degree of frequency (from never/rarely, sometimes, always) compared to physicians not working in such units. Physicians working in the North and Central area of the country, the most affected regions, also reported higher odds of having adequate access to PPE (OR=2, 95% C.I. 1.4–3) compared to those working in the South. On the contrary, adult primary care physicians had half the odds (OR=0.5, 95% C.I. 0.3–1) of having access to PPE when they needed it. See Table 2.

**Table 2.** Predictors of physicians’ access to PPE, information about the use of PPE, donning and doffing performance, and risk perception at work.

| Independent variables | Multiple regression models - Odds ratios and 95% confidence limits |  |  |  |
| --- | --- | --- | --- | --- |
|  | Access to PPE* | Information received about the use of PPE* | Ability to perform donning and doffing procedures** | Risk perception of contracting COVID-19 at work* |
| Years of work experience (<5;5-10;11-20;21-30;>30) | N/A | 1.1 [1-1.3] | 1.5 [1.3-1.8] | N/A |
| Working in a COVID-19 unit (Yes=1; No=0) | 3.8 [2.5-5.7] | 1.8 [1.2-2.5] | 2.4 [1.5-3.7] | 2.4 [1.7-3.6] |
| Region (North/Central=1; South=0) | 2 [1.4-3] | N/A | 1.3 [0.8-2.1] | N/A |
| Working as an adult primary care physician (Yes=1; No=0) | 0.5 [0.3-1] | 0.6 [0.3-1] | 0.5 [0.2-1] | 2.4 [1.7-3.6] |
| Receiving adequate information about the use of PPE (Never/Rarely=1; 2=Sometimes; 3=Always) | N/A | N/A | 2.2 [1.7-2.8] | 0.5 [0.4-0.6] |

\*ordered logistic regression;
\*\* logistic regression
N/A = excluded after bivariate analysis

### Information about the use of PPE

When physicians were asked how frequently they had *received adequate information regarding the use of PPE to protect themselves from contracting COVID-19*, 136 (26%) reported that they always did, 198 (37%) sometimes, 195 (36%) rarely or never. Approximately half of the physicians reported that the information received to date about the use of PPE was either clear (47%) or complete (54%) and approximately one quarter was unsure about clarity (28%) or completeness (27%), leaving only 25% satisfied with the information they received. When asked if the information received was useful to them, opinions were equally split between three groups: those who found it useful (33%), those who did not (35%), and those who were unsure about its usefulness (31%). As a result of the bivariate analysis years of experience, working in a COVID-19 unit and in a primary care setting were associated to the dependent variable, while geographic area (North, Centre, or South) was dropped from the final ordered logistic multiple model because of p-value > 0.25. In the ordered logistic multiple regression model for each increase in the category of years of experience (rarely or never, sometimes, always) there was a 1.1 increased odds (OR=1.1, 95% C.I. 1–1.3) of having received *adequate information about the use of PPE* at a higher degree of frequency (from never/rarely, sometimes, always). Physicians working in a unit dedicated to COVID-19 also reported higher odds (OR=1.76, 95% C.I. 1.2–2.5) of receiving adequate information more frequently compared to physicians not working in such units. On the contrary, adult primary care physicians had 0.6 decreased odds (OR=0.6, 95% C.I. 1.2–2.5) of receiving such information compared to physicians working in a different setting (hospital or pediatric primary care). See Table 2.

### Ability to perform donning and doffing procedures

When asked if they believed they could correctly execute donning and doffing procedures for specific pieces of PPE, respondents felt mostly unprepared for putting the respirators and gowns on (14% and 12% respectively) or unsure if they were doing it correctly (12% and 25%). In regards to doffing, once again, taking off the respirator and the gown were the procedures they did not know how to do correctly (19% and 34%) or were unsure about (34% and 31%). See Figure 1. In the multiple logistic regression model years of experience, working in a covid unit, and frequency of information received about the use of PPE were associated with better donning and doffing performance. For each increase in the category of years of experience physicians reported 1.5 greater odds of being able to perform the procedures [OR=1.5, 95% C.I. 1.3–1.8], those working in COVID-19 units have twice the odds of reporting being able to correctly perform the procedures [OR=2.3, 95% C.I. 1.5–3.7], and finally those who reported to have received adequate information during the epidemic also reported greater odds of being able to perform the procedures [OR=2.2, 95% C.I. 1.7–2.7]. See Table 2.

**Figure 1.**
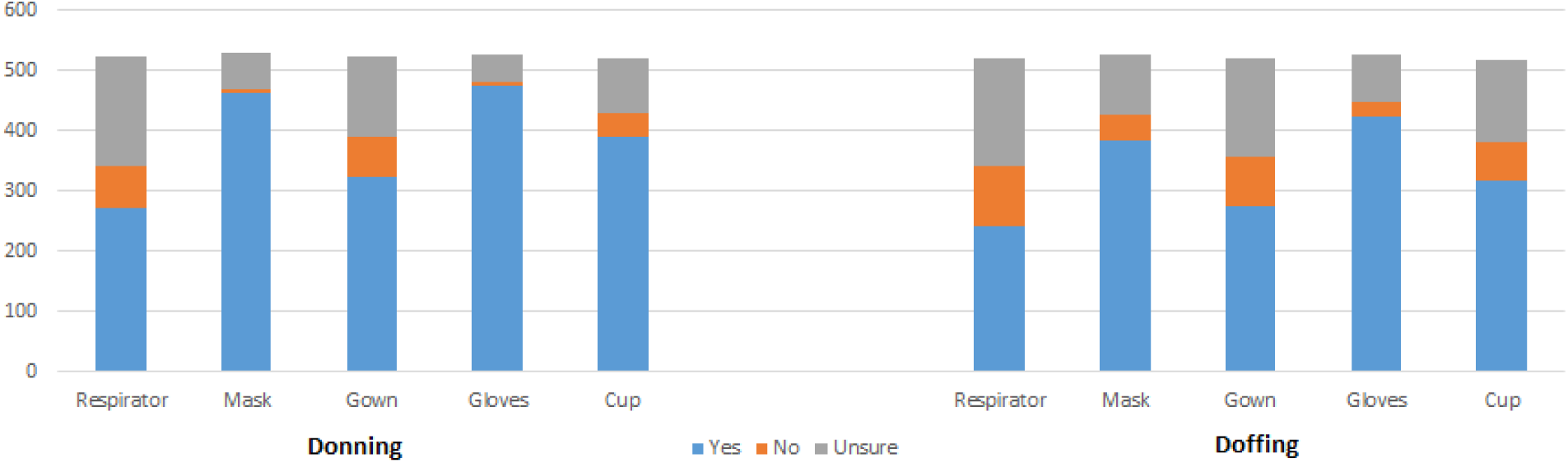
Based on the information received to date do you believe you can correctly execute donning and doffing procedures for the following PPE?

### Accuracy of PPE knowledge based on current guidance

As part of the survey we also presented the physicians with 12 scenarios of activities that would require the use of different types of PPE and asked them, based on their knowledge, what was the most appropriate piece of equipment for each activity. The activities included transportation of presumptive and positive COVID-19 patients within the hospital or by ambulance, activities in the triage area, routine physical examination of patients with respiratory and without respiratory symptoms and administrative activities with direct contact with clients. Overall respondents assigned to each activity a level of protection higher compared to what is currently recommended by current guidance.^10^ For example over 90% of physicians said that a face shield is appropriate when conducting physical examination on a COVID-19 positive patient while a surgical mask is what is typically recommended. Similarly over 70% physicians reported that a face shield is needed when conducting the same routine examination in any patient with respiratory symptoms. Interestingly gloves were reported as appropriate by over 75% of physicians for the examination of any patient even if without respiratory symptoms. And over 70% reported as appropriate the use of a surgical mask when performing administrative duties. See Table 2.

### Risk perception

When physicians were asked to rate their perceived risk, on a scale from 0 to 100, of contracting the infection in the healthcare setting, they attributed a mean value of 56 (SD=22) to such risk, the same perception of risk for their life outside the work environment was much lower 25 (SD=19) T test p<0.001. Though physicians perceived the risk of contracting COVID-19 outside of work to be less, they were still concerned about infecting their family members upon return home from work. The majority of the respondents (73%) reported to have taken precautions at home to keep their family safe; examples consisted in removing and washing their clothes upon arrival at home and wearing a surgical mask, sleeping in a separate room, not sharing utensils and keeping a physical distance from family members. Interestingly, none reported to check their body temperature at home. In an attempt to strengthen their immune system and prevent the infection, despite lack of evidence on the matter 35% of respondents, reported to have taken vitamins (mainly C and D) and (1%) self-administered hydroxicloroquine. In the multivariable ordered logistic model of factors related to perceived risk at work, those working in COVID-19 units showed 3.2 greater odds of reporting a higher level of risk perception compared to those not working in such units [OR=3.2, 95% C.I. 2.1–4.7]. On the contrary those with access to PPE showed lower odds of reporting higher risk perception [OR=0.6, 95% C.I 0.4–0.7] compared to those who do not to have access to the equipment, and so do those who report to have received adequate information on how to use the PPE [OR=0.6, 95% C.I. 0.4–0.7]. Interestingly, the ability to conduct donning or doffing procedures was not associated with risk perception. See Table 2. Interestingly, among all specialties physicians in dentistry, otolaryngology, occupational medicine and pneumology were the ones that showed the highest risk perception based on the descriptive analysis. The lowest level of risk perception was reported by those holding a specialty in hygiene and preventive medicine which is consistent with the fact that these professionals are typically in healthcare management positions and do not directly assist patients. See Figure 2.

**Figure 2.**
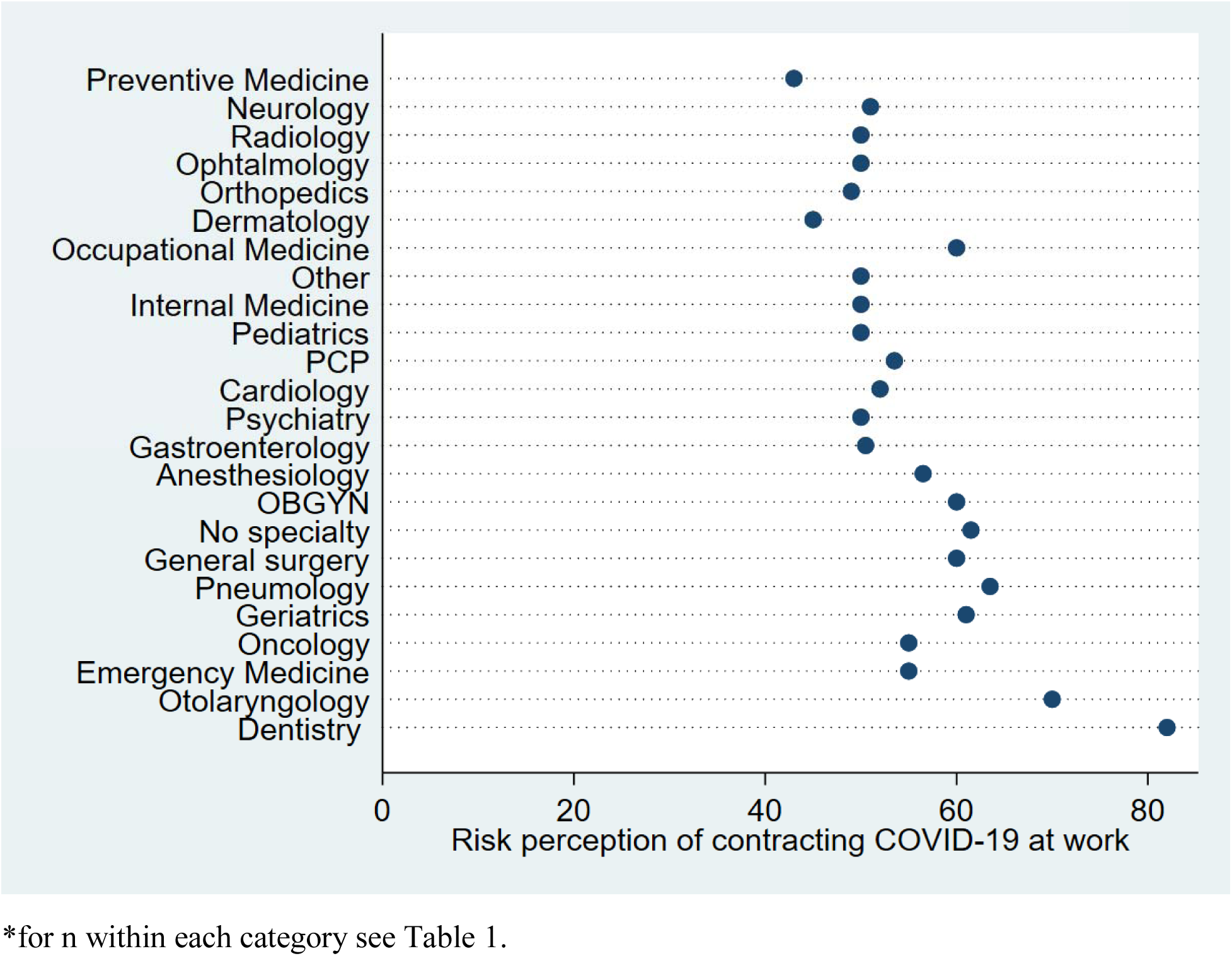
Risk perception by specialty*

## Discussion

Our results present some of the first evidence on how Italian physicians experienced lack of PPE, and what factors influenced their understanding of PPE procedures and use. While our results are not generalizable to the all population of Italian physicians, and certainly derived from a group of physicians with high level of interest in COVID-19, group differences within our sample rather than general group estimates by extrapolation, can be useful to understand predictors of behaviors and specific challenges in access and use of PPE. The majority of those surveyed reported not to have access to PPE every time they need it and at least one third of them reported not having received adequate information on the use of the equipment, nor were they consistently comfortable with donning and doffing procedures, in particular when using respirators and wearing gowns. Working in a COVID unit made a difference in multivariate analysis of both having access to PPE, adequate information on their use, feeling comfortable with donning and doffing procedures, and perceived risk. This likely reflects training efforts focused on educating this subset of the workforce, those actually at the highest risk of contracting COVID-19 based on occupational risk. However, given the difficulties of creating 100% COVID-19 free clinics as many patients may present to a clinic in a pauci-symptomatic status, the current variation in access and knowledge about PPE use, may put at a disproportionate risk those working outside COVID-19 units. More specifically, our results indicate how primary care physicians may have been neglected from informational initiatives posing them at high risk of contracting the infection. With the advancement of testing and treatment options in the months ahead more and more COVID-19 patients will be diagnosed and cured outside the hospital setting. Therefore, additional attention is needed to provide PPE and PPE training for this group of providers and all those working outside COVID-19 units.

Interestingly, respondents consistently overestimated the level of PPE needed to interact with a non-symptomatic patient; reflecting they either had inadequate understanding of current guidance or regardless of the guidance they were fearful of becoming infected themselves and/or infecting the patient when a diagnosis was not confirmed. The ongoing changes to PPE guidance provided by international, national and regional public health agencies, in particular in regards to the use of respirators, likely made it more challenging to make sense of which equipment to use. Standards of use evolved in mid-March, a couple of weeks prior to our survey, as a result of PPE shortages and lack of logistics planning within hospitals. Limited training as well as pre-existing professional norms that lacked a culture of PPE use may have been factors that shaped challenges in developing adequate training and information material. We suggest that future efforts should be made to include PPE training in the medical curriculum so that in times of crisis physicians can better adapt to their use and differences in knowledge and practices would be less evident across categories. Methods for just-in-time training including the use of video trainings may be one mechanism to improve donning and doffing procedures.^11^ In times of crisis, an overuse and gauging of PPE by concerned physicians may cause as much harm as lack of supplies.

We found PPE perceptions and use were also tied to perceived risk of contracting the infection in the work environment. Overall, risk perception was high, but both adequate access and PPE training decreased such perception. Of concern, is also the fact that many physicians took actions in their personal lives to protect their families, limiting physical interactions and in some cases renting separate apartments. In the long term, these actions will certainly affect their emotional well being.

## Conclusion

Results from this rapid survey indicate that while ramping up supplies on PPE for healthcare workers is a necessity, adequate training and clear instructions are just as important. To the extent possible instructions need to be consistent overtime and across regions, include recommendations not only on the overall safety of the workers in the healthcare setting but also on strategies to maintain their overall physical and emotional health and the health of their loved ones.

## Acknowledgments

We would like to thank for their support the administrator and all of the moderators of the Facebook private group “Coronavirus, Sars-CoV-2 e COVID-19 gruppo per soli medici - https://www.facebook.com/groups/Coronavirusmediciitaliani/” We are also grateful to all of the colleagues of the group which decided to compile the questionnaire giving their suggestions and point of view on this topic”.

## Data Availability

Data is available upon request from the authors.

